# Solicited Cough Sound Analysis for Tuberculosis Triage Testing: The CODA TB DREAM Challenge Dataset

**DOI:** 10.1101/2024.03.27.24304980

**Authors:** Sophie Huddart, Vijay Yadav, Solveig K. Sieberts, Larson Omberg, Mihaja Raberahona, Rivo Rakotoarivelo, Issa N. Lyimo, Omar Lweno, Devasahayam J Christopher, Nguyen Viet Nhung, Grant Theron, William Worodria, Charles Y. Yu, Christine M Bachman, Stephen Burkot, Puneet Dewan, Sourabh Kulhare, Peter M Small, Adithya Cattamanchi, Devan Jaganath, Simon Grandjean Lapierre

**Author notes:** CORRESPONDING AUTHOR: Simon Grandjean Lapierre. Equal contributions (Sophie Huddart, Vijay Yadav, Devan Jaganath, Simon Grandjean Lapierre).

## Abstract

Cough is a common and commonly ignored symptom of lung disease. Cough is often perceived as difficult to quantify, frequently self-limiting, and non-specific. However, cough has a central role in the clinical detection of many lung diseases including tuberculosis (TB), which remains the leading infectious disease killer worldwide. TB screening currently relies on self-reported cough which fails to meet the World Health Organization (WHO) accuracy targets for a TB triage test. Artificial intelligence (AI) models based on cough sound have been developed for several respiratory conditions, with limited work being done in TB. To support the development of an accurate, point-of-care cough-based triage tool for TB, we have compiled a large multi-country database of cough sounds from individuals being evaluated for TB. The dataset includes more than 700,000 cough sounds from 2,143 individuals with detailed demographic, clinical and microbiologic diagnostic information. We aim to empower researchers in the development of cough sound analysis models to improve TB diagnosis, where innovative approaches are critically needed to end this long-standing pandemic.

## BACKGROUND AND SUMMARY

Tuberculosis remains the leading infectious disease killer globally, partly due to public health systems’ inability to accurately diagnose millions of infected individuals every year.^1^ Insufficient access to high-quality TB screening and diagnosis is recognized as one of the most important gaps in the cascade of care.^2^ Here we describe a cough sound database including detailed demographic, clinical and microbiologic information for the development of AI-based sound classification TB triage models. As the WHO’s End TB Strategy calls for intensified research and innovation including the discovery of new tools for community-based screening, digital cough monitoring and *Acoustic Epidemiology* could represent new tools that can help bend the TB pandemic curve and accelerate the achievement of global TB elimination goals.^3–5^

The “missing millions” of undiagnosed patients living with active TB disease represent an heterogeneous group including those who did not access triage or diagnosis testing or weren’t appropriately referred for effective treatment. Improving the accuracy, portability, point-of-care amenability and connectivity of diagnostic tools and algorithms would have significant value. Most health systems build their TB programs on a combination of complementary screening followed by diagnostic tests. The WHO’s target product profile (TPP) for a community-based TB triage test suggests that it should be at least 90% sensitive and 70% specific.^6^ According to the 2021 WHO TB screening guidelines, symptom-based screening with questionnaires, including cough, is 42% sensitive.^7^ Besides having poor accuracy these guidelines have operational challenges that impede its sustained and uniform implementation within resource-challenged TB programs. Other tools such as digital chest X-rays combined with computer-aided detection (CAD) algorithms have also been evaluated in the context of TB triage. This approach was shown to be highly sensitive but had variable specificity and remains difficult to deploy due to limited availability of chest X-ray platforms at primary-level health facilities.^8^ Whether in the context of community-based outreach screening or healthcare facility-based evaluation prior to confirmatory testing, cough classification models could complement or replace other triage strategies including symptom-based screening.

We historically have been unable to objectively monitor cough sounds and consequently reduced this data-rich symptom into subjective and dichotomous information (e.g., cough versus no cough, chronic versus acute, better versus worse). Advances in acoustics and machine learning (ML) have enabled the identification and recording of human coughs in real-world acoustic environments (cough detection) as well as differentiation of coughs from patients with distinct clinical conditions or at different stages of disease (cough classification). As part of the emerging field of *Acoustic Epidemiology,* this has the potential to develop novel screening or diagnostic assays with simple digital recording devices, such as a smartphone, tablet or watch.^5^ Proof-of-concept studies previously showed that cough associated with TB contains a specific acoustic signature which can be recognized by ML models. A study by *Pahar et al*. suggests that a cough-based TB screening model can discriminate TB cough sounds from those associated with other lung conditions with 93% sensitivity and 95% specificity, exceeding the WHO TPPs.^9^ In a study combining cough sound analysis and patients’ clinical characteristics, *Yellapu et al.* report that ML can be used to detect TB with 90% sensitivity and 85% specificity.^10^ Those pilot studies report on ML models which were designed on small datasets and were not validated in external populations. Given the potential impact on performance of local disease epidemiology and population ethnicity among other confounders, large and diverse cough datasets are needed to replicate those studies.

We collected and are here releasing a dataset including 733,756 cough sounds from 2,143 patients across 7 countries with accurately annotated demographic, clinical and microbiologic diagnostic information. These data were initially used to enable and evaluate the CODA TB DREAM Challenge which invited participants to develop algorithms for prediction of TB diagnosis. The training data are now available for general use, and researchers are invited to leverage acoustic and clinical data to further develop and evaluate sound classification models for TB screening against a held-out test partition.^11^ We aim to enable the development of models which could achieve the WHO TPP performance targets for the current ‘community-based TB triage test’ or the forthcoming TPP for a TB screening test.^6,12^ This data set has limitations which include some selection bias since it was collected from a symptomatic presumptive TB population. The developed models which will be developed may hence not perform as well if used for asymptomatic screening at population level. Accordingly, more data should be collected from community screening activities.

## METHODS

### Participants

A total of 2,143 participants were recruited from two parent studies described below. To be eligible, participants had to be 18 years or older and have a new or worsening cough for at least two weeks. took place at outpatient clinics in India, Madagascar, the Philippines, South Africa, Tanzania, Uganda, and Vietnam. All participants provided informed consent. A summary of participant demographics and country distribution are available in Table 1.

**Table 1.** Participant demographics across training and test sets.

|  | **Training set**<br>**(N=1105)** | **Testing set**<br>**(N=1038)** | **Complete**<br>**set**<br>**(N=2143)** |
| --- | --- | --- | --- |
| **Sex** |  |  |  |
| Female | 517 (46.8%) | 460 (44.3%) | 977 (45.6%) |
| Male | 588 (53.2%) | 578 (55.7%) | 1166 (54.4%) |
| **Age** |  |  |  |
| Median [Q1, Q3] | 40.0 [28, 53] | 40.0 [29, 53] | 40.0 [28, 53] |
| **Height (cm)** |  |  |  |
| Median [Q1, Q3] | 162 [155, 168] | 162 [156, 168] | 162 [156, 168] |
| **Weight (Kg)** |  |  |  |
| Median [Q1, Q3] | 55.0 [49, 65] | 56.0 [49, 65] | 55.8 [49, 65] |
| **HIV status** |  |  |  |
| Negative | 878 (79.5%) | 809 (77.9%) | 1687 (78.7%) |
| Positive | 162 (14.7%) | 155 (14.9%) | 317 (14.8%) |
| Unknown | 65 (5.9%) | 74 (7.1%) | 139 (6.5%) |
| **Duration of cough (days)** |  |  |  |
| Median [Q1, Q3] | 30.0 [16, 60] | 30.0 [16, 50] | 30.0 [16, 60] |
| **Prior TB** |  |  |  |
| No | 903 (81.7%) | 835 (80.4%) | 1738 (81.1%) |
| Not sure | 3 (0.3%) | 2 (0.2%) | 5 (0.2%) |
| Yes | 199 (18.0%) | 201 (19.4%) | 400 (18.7%) |
| **Country** |  |  |  |
| India | 119 (10.8%) | 122 (11.8%) | 241 (11.2%) |
| Madagascar | 159 (14.4%) | 75 (7.2%) | 234 (10.9%) |
| The Philippines | 198 (17.9%) | 190 (18.3%) | 388 (18.1%) |
| South Africa | 137 (12.4%) | 138 (13.3%) | 275 (12.8%) |
| Tanzania | 87 (7.9%) | 110 (10.6%) | 197 (9.2%) |
| Uganda | 242 (21.9%) | 245 (23.6%) | 487 (22.7%) |
| Vietnam | 163 (14.8%) | 158 (15.2%) | 321 (15.0%) |
| **Microbiologic reference standard** |  |  |  |
| TB Negative | 807 (73.0%) | 782 (75.3%) | 1589 (74.1%) |
| TB Positive | 297 (26.9%) | 256 (24.7%) | 553 (25.8%) |
| **Sputum Xpert reference standard** |  |  |  |
| Indeterminate | 4 (0.4%) | 2 (0.2%) | 6 (0.3%) |
| TB Negative | 839 (75.9%) | 808 (77.8%) | 1647 (76.9%) |
| TB Positive | 262 (23.7%) | 226 (21.8%) | 488 (22.8%) |

#### Rapid Research in Diagnostic Development TB Network (R2D2 TB Network) study

The R2D2 TB Network study evaluates novel TB diagnostics in various stages of development among people with presumptive TB in five low- and middle-income countries: Uganda, South Africa, Vietnam, the Philippines and India.^13^ Ethical approval for this study was obtained from institutional review boards (IRB) in the US and in each study site. In the US, approval was obtained from the University of California San Francisco IRB (# 20-32670). In Vietnam, approval was obtained from the Ministry of Health Ethical Committee for National Biological Medical Research (94/CN-HĐĐĐ), the National Lung Hospital Ethical Committee for Biological Medical Research (566/2020/NCKH) and the Hanoi Department of Health, Hanoi Lung Hospital Science and Technology Initiative Committee (22/BVPHN). In India, approval was obtained from Christian Medical College IRB (13256). In South Africa, approval was obtained from Stellenbosch University Health Research Ethics Committee (17047). In Uganda, approval was obtained from Makerere University, College of Health Sciences, School of Medicine, Research Ethics Committee (2020-182). In the Philippines, approval was obtained from De La Salle Health Sciences Institute Independent Ethics Committee (2020-33-02-A).

#### The Digital Cough Monitoring for screening, diagnosis and clinical follow-up of tuberculosis and other respiratory diseases project

This project was designed to embed digital cough monitoring within existing health facility-based TB diagnostic cohorts in Madagascar and Tanzania. Ethical approval for this study was obtained from institutional review boards (IRB) in Canada and in each study site. In Canada, approval was obtained from the Centre de Recherche du Centre Hospitalier de l’Université de Montréal IRB (# 2021-9270, 20.226). In Madagascar, approval was obtained from the Comité d’Éthique à la Recherche Biomédicale (IORG0000851 - N°051-MSANP/SG/AMM/CERBM).). In Tanzania, approval was obtained from the Ifakarah Health Institute IRB (31-2021) and the National Institute for Medical Research (NIMR/HQ/R.8a/Vol IX/3805).

### Data Collection

#### Demographic and clinical data

At enrollment into the parent studies, participants underwent a baseline questionnaire, clinical examination, and sputum collection for TB testing. Study staff also recorded participants’ age, gender, height, weight, smoking status and duration of cough. HIV diagnosis was made either based on participant self-report of a positive HIV diagnosis or a positive test result. A summary of the available variables is shown in Table 2.

**Table 2.** Available demographic, clinical and microbiologic variables.

| Variable | Values | Format/Definition |
| --- | --- | --- |
| Country | Philippines<br>Vietnam<br>South Africa<br>Uganda<br>India<br>Madagascar<br>Tanzania | Country where the participant was enrolled |
| Sex | Male<br>Female | Sex at birth reported by participant |
| Age | Numeric | Age calculated as date of collection - date of birth if known. If date of birth is unknown, reported age at time of collection. |
| Height | Numeric | Height in centimeters |
| Weight | Numeric | Weight in Kg |
| HIV status | Positive<br>Negative | HIV status. All participants who do not report being HIV-positive receive HIV testing (using capillary or venous blood)<br>Positive = Positive reported by patient or positive on a HIV test<br>Negative = Negative on a HIV test |
| Reported duration of cough | Numeric | Self-reported duration of current cough (days). At baseline, we ask: How many days have you had this new cough or cough that has been worse? |
| Prior TB | Yes<br>No | Self-reported. At baseline, we ask: Have you ever had or been told you had tuberculosis (TB)? |
| Prior TB type: Pulmonary | Checked<br>Unchecked | Asked at baseline: "With what kind of TB were you diagnosed?" (may select more than one)<br>Participant selected pulmonary TB. |
| Prior TB type: Extrapulmonary | Checked<br>Unchecked | Asked at baseline: "With what kind of TB were you diagnosed?" (may select more than one)<br>Participant selected extrapulmonary TB. |
| Prior TB type: Unknown | Checked<br>Unchecked | Asked at baseline: "With what kind of TB were you diagnosed?" (may select more than one)<br>Participant selected Unknown. |
| Hemoptysis | Yes<br>No | In the past 30 days, have you ever coughed up blood? |
| Heart rate | Numeric | Participant's heartrate (beats per minute) measured at baseline |
| Temperature | Numeric | Participant's temperature (Celsius) measured at baseline. |
| Weight loss | Yes<br>No | Subjective, Self-reported. At baseline, we ask: In the past 30 days, have you experienced any weight loss? |
| Smoked in last week | Yes<br>No | Asked at baseline "Have you used combustible tobacco and/or vaping products in the last 7 days?" |
| Fever | Yes<br>No | Self-reported fever at baseline: "In the past 30 days, have you ever felt or experienced fever?" |
| Night sweats | Yes<br>No | Self-reported night sweats at baseline: "In the past 30 days, have you ever experienced night sweats?" |
| Microbiologic reference standard* | TB Positive<br>TB Negative<br>Indeterminate | TB diagnosis based on sputum and culture results |
| Sputum Xpert reference standard* | Positive<br>Negative<br>Indeterminate | TB diagnosis based on sputum results alone |
| Xpert combined semi-quant | Trace<br>Very Low<br>Low<br>Medium<br>High | Highest semiquantitative result from sputum Xpert Ultra test conducted at baseline visit. A semiquantitative result will only be available if the test was positive. |

#### TB Reference Standard Testing

Both Xpert MTB/RIF Ultra PCR and mycobacterial culture (Lowenstein-Jensen solid medium or MGIT liquid medium) were performed on sputum collected from all participants. Any participant whose first sputum Xpert MTB/RIF Ultra result was indeterminate or trace-positive, received a second sputum Xpert MTB/RIF Ultra test. Results from those assays were combined to determine TB status according to two reference standards: a microbiologic reference standard and a sputum Xpert reference standard. The sputum Xpert reference standard is restricted to Xpert MTB/RIF Ultra results on sputum samples. The microbiologic reference standard includes culture results, allowing for more individuals to be classified as TB positive. The microbiologic reference standard is considered the primary reference standard. Full details of the reference standards are described in Table 3.

**Table 3.**
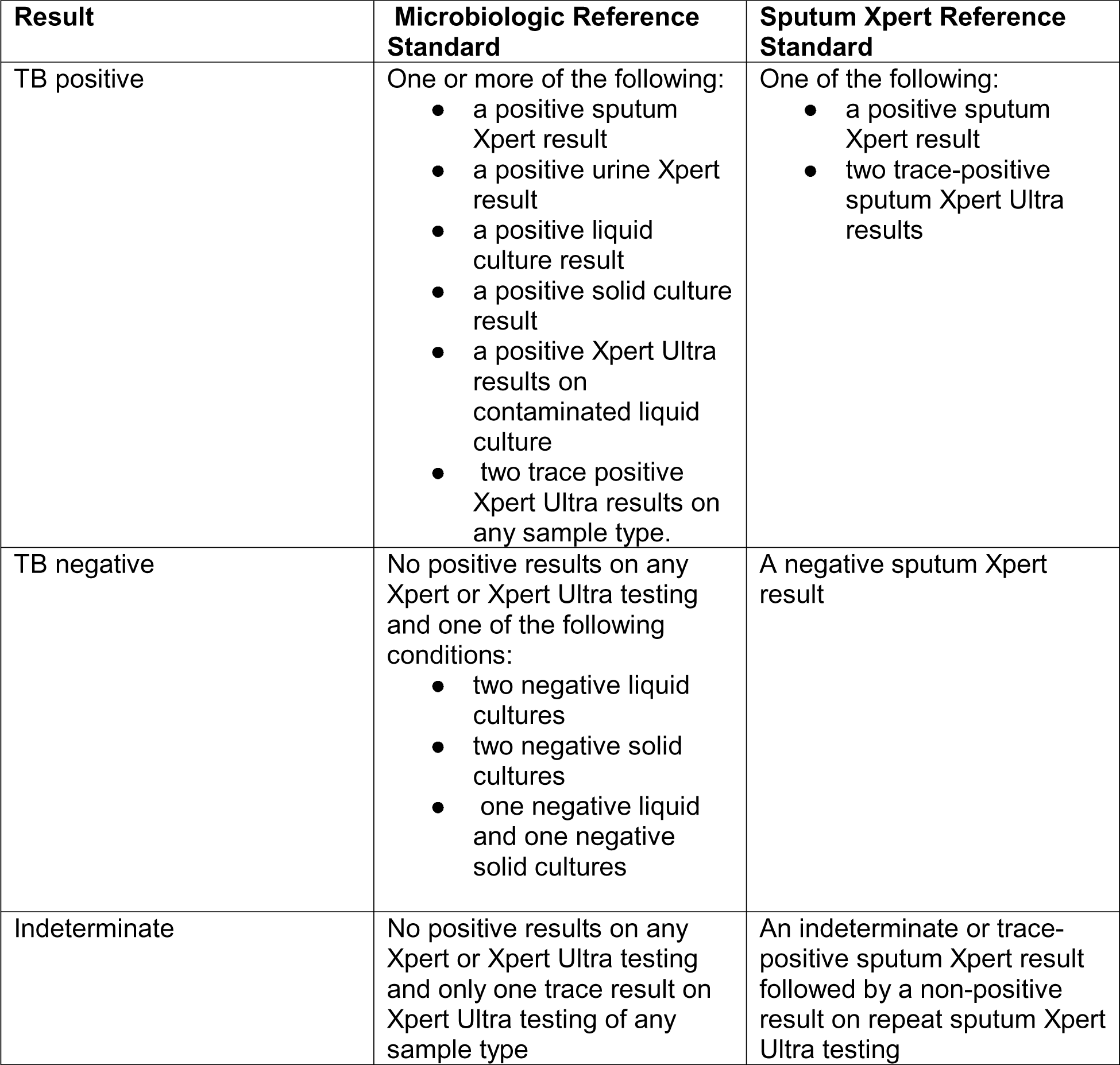
Reference standard definitions.

| **Result** | **Microbiologic Reference Standard** | **Sputum Xpert Reference Standard** |
| --- | --- | --- |
| TB positive | One or more of the following:<br>• a positive sputum Xpert result<br>• a positive urine Xpert result<br>• a positive liquid culture result<br>• a positive solid culture result<br>• a positive Xpert Ultra results on contaminated liquid culture<br>• two trace positive Xpert Ultra results on any sample type. | One of the following:<br>• a positive sputum Xpert result<br>• two trace-positive sputum Xpert Ultra results |
| TB negative | No positive results on any Xpert or Xpert Ultra testing and one of the following conditions:<br>• two negative liquid cultures<br>• two negative solid cultures<br>• one negative liquid and one negative solid cultures | A negative sputum Xpert result |
| Indeterminate | No positive results on any Xpert or Xpert Ultra testing and only one trace result on Xpert Ultra testing of any sample type | An indeterminate or trace-positive sputum Xpert result followed by a non-positive result on repeat sputum Xpert Ultra testing |

#### Cough Recording

Cough sounds were collected using smartphones loaded with the Hyfe research app.^14^ Specific phone models used in the different participating sites are presented in Supplementary Materials 1. Hyfe research app is designed to listen for explosive sounds and record *∼*0.5 seconds sound fragments corresponding to putative cough sounds. Hyfe research app uses a server-based convolutional neural network (CNN) model to classify explosive sounds as coughs and recordings of these cough sounds are saved on a protected health information (PHI)-regulated server for analysis. This model has been shown to be 96% sensitive and 96% specific for cough detection using human-labeled sounds as a reference standard.^15^ Smartphones were positioned on tripods in rooms within the clinic. Participants were asked to cough five times (solicited cough) while standing 60-90 cm from the tripod; participants who managed to produce at least three coughs were retained in the dataset. Some participants produced more than five coughs due to a triggered coughing fit and those additional coughs were also collected and included in the dataset. Solicited and triggered coughs could not be labeled distinctively and are treated the same in the dataset. After enrollment and onboarding, a subset of participants (n = 565) were also asked to carry a study phone for two weeks and collect longitudinal coughs sounds in an outpatient setting. Those sounds are labeled as longitudinal and made available within the dataset. A tally of solicited and longitudinal cough sounds per data partition are available in Table 4.

**Table 4.**
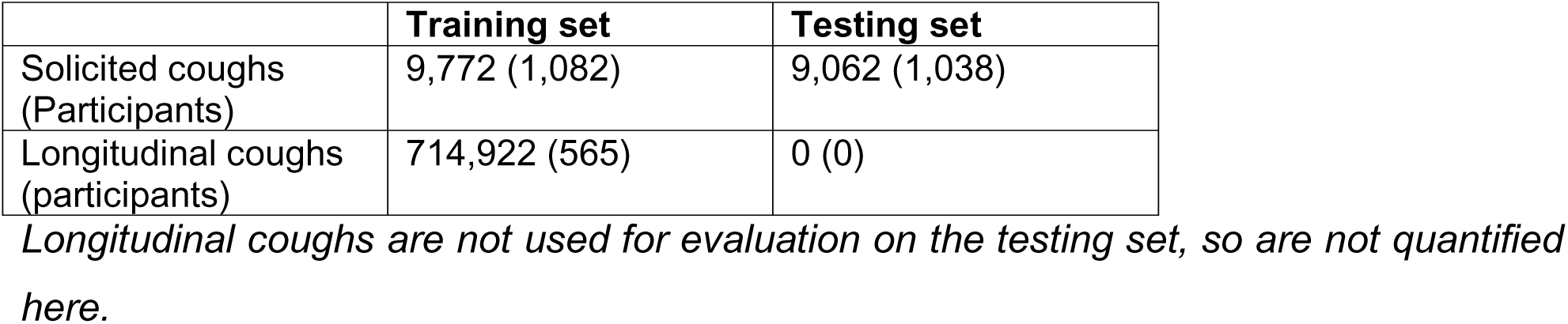
Number of cough recordings per data partition.

|  | **Training set** | **Testing set** |
| --- | --- | --- |
| Solicited coughs (Participants) | 9,772 (1,082) | 9,062 (1,038) |
| Longitudinal coughs (participants) | 714,922 (565) | 0 (0) |

*Longitudinal coughs are not used for evaluation on the testing set, so are not quantified* *here.*

### Data Partitioning

The dataset was split into a training (n=1,105) and validation set (n=1,038). The dataset was randomly partitioned evenly between the training and testing set at the level of the participant (i.e., all of a participant’s cough sounds are in either the training or validation set).

### Data Pre-Processing

#### Cough sounds

The sound recordings available in this dataset have not undergone pre-processing beyond their identification as a cough sound by the Hyfe research app CNN model.

#### Clinical Data

Data from all participating sites were collected with standardized questionnaires and definitions. Data formatting was harmonized in the open access database.

### Dataset Description

Sage Bionetworks independently verified the variable balance between the training and validation sets as demonstrated in Table 1. A breakdown of key demographics and microbiologic reference standard results by country are shown in Table 5.

**Table 5.** Key variables summarized by country.

|  | **India**<br>n = 241 | **Madagascar**<br>n = 234 | **Philippines**<br>n = 388 | **South Africa**<br>n = 275 | **Tanzania**<br>n = 197 | **Uganda**<br>n = 487 | **Vietnam**<br>n = 321 |
| --- | --- | --- | --- | --- | --- | --- | --- |
| **Sex** |  |  |  |  |  |  |  |
| Female | 100 (41%) | 115 (49%) | 208 (54%) | 144 (52%) | 86 (44%) | 203 (42%) | 121 (38%) |
| Male | 141 (59%) | 119 (51%) | 180 (46%) | 131 (48%) | 111 (56%) | 284 (58%) | 200 (62%) |
| **Age** |  |  |  |  |  |  |  |
| Median [Q1, Q3] | 47 [34, 58] | 32 [24, 51] | 39 [26, 53] | 40 [32, 49] | 40 [31, 50] | 32 [26, 42] | 54 [41, 64] |
| **Duration of cough (days)** |  |  |  |  |  |  |  |
| Median [Q1, Q3] | 45 [30, 90] | 30 [17, 60] | 21 [15, 30] | 21 [14, 28] | 30 [14, 30] | 30 [16, 60] | 30 [30, 90] |
| **Prior TB** |  |  |  |  |  |  |  |
| No | 204 (85%) | 207 (88%) | 325 (84%) | 170 (62%) | 147 (75%) | 427 (88%) | 258 (80%) |
| Not sure | 0 (0%) | 0 (0%) | 4 (1.0%) | 0 (0%) | 0 (0%) | 0 (0%) | 1 (0.3%) |
| Yes | 37 (15%) | 27 (12%) | 59 (15%) | 105 (38%) | 50 (25%) | 60 (12%) | 62 (19%) |
| **Microbiologic reference standard** |  |  |  |  |  |  |  |
| TB Negative | 216 (90%) | 131 (56%) | 348 (90%) | 223 (81%) | 161 (82%) | 313 (64%) | 199 (62%) |
| TB Positive | 25 (10%) | 103 (44%) | 40 (10%) | 52 (19%) | 36 (18%) | 174 (36%) | 122 (38%) |

## DATA RECORDS

De-identified participant demographic and clinical data, including TB reference standard results, cough sound WAV files, and a datafile linking participant IDs to sound file IDs were exported to a dedicated project in Synapse. Synapse is a general-purpose data and analysis sharing service where members can work collaboratively, analyze data, share insights, and have attributions and provenance of those insights to share with others. Synapse is developed and operated by Sage Bionetworks^16^. A total of 1,105 participants’ data are made available for access and download as a training dataset. The validation set is withheld, but models can be evaluated against the validation set via the instructions provided in the Synapse project.

All training set files are stored and are accessible via the Synapse platform with associated metadata and documentation and can be accessed at the following URL: www.synapse.org/TBcough - https://doi.org/10.7303/syn31472953.

## TECHNICAL VALIDATION

All cough collection periods were observed by study staff and cough sounds were spot-checked for accurate recording. Patient metadata was reviewed by study staff for accuracy. The data described in this article were collected using the Hyfe Research app which uses a proprietary algorithm to identify cough sounds. We used a prediction score of 0.8 from this algorithm to filter potential non-cough sounds. To validate the precision of the Hyfe algorithm, a standalone computer vision and deep learning model was trained using Log Mel spectrogram images from ESC-50 and Coswara datasets.^17,18^ The “VGG16” CNN based pre-trained model was trained for accurate classification of cough sounds and achieved a model accuracy of approximately 96% on Hyfe cough recordings.^19^ Most of the recordings that were classified incorrectly had a Hyfe prediction score less than 0.8.

## USAGE NOTES

Users can register to evaluate predictive models of TB diagnosis against the held-out test partition via the instructions on the Synapse project. The scoring mechanism can evaluate two different types of models: (1) those that use only cough sounds, or (2) those which also incorporate clinical metadata variables which have been provided in the training dataset (sex, age, height, weight, reported duration of cough, prior TB diagnosis and type, hemoptysis, heart rate, temperature, weight loss, smoking in the last week, fever and night sweats). Models are submitted to the scoring queues as Docker images. Full instructions and example code is available on Synapse project website (www.synapse.org/TBcough).

### Downloading the Data

Given the number of files represented in the data, users should consider downloading the data via one of the programmatic Synapse clients (available in R or Python). For convenience, Python code for downloading the data is provided in the Synapse project wiki. The training dataset size is (0.43 GB) for the solicited coughs and (31.6 GB) for the longitudinal coughs.

### Data Use Agreement

To access the data, individuals must become Certified and Validated users of Synapse and maintain an active account on Synapse: http://www.synapse.org. They must also submit an Intended Data Use Statement and agree to the Terms of Use of the dataset. Terms of Use are summarized in Supplementary Materials 2.

## CODE AVAILABILITY

No additional data processing was conducted other than what has been described above.

## DATA AVAILABILITY

All training set files are stored and are accessible via the Synapse platform with associated metadata and documentation and can be accessed at the following URL: www.synapse.org/TBcough.

Specific acoustic and clinical metadata as well as dataset information can be found under the following doi references.

Clinical data: https://doi.org/10.7303/syn53710097

Cough metadata: https://doi.org/10.7303/syn53710098

Solicited Coughs: https://doi.org/10.7303/syn40358494

Longitudinal Coughs: https://doi.org/10.7303/syn40358476

Data Dictionary: https://doi.org/10.7303/syn41743692

Data sharing and model benchmarking website: https://doi.org/10.7303/syn31472953

## SUPPLEMENTARY MATERIALS

### Supplementary materials 1 Phone models used in the different participating sites

India

- Redmi 9 Prime
- Realme Narzo20

Madagascar

- Motorola G9 play

Philippines

- Myphone myWX2 Pro
- Xiaomi 9C

South Africa

- Nokia 3.1
- Nokia 5.4
- Xiaomi Redmi 9A

Tanzania

- Nokia 3.4 Ta-1288

Uganda

- Motorola G16
- Samsung M11
- Nokia model 5.3

Vietnam

- OPPOA54

### Supplementary materials 2 Data Use Agreement

Researchers wishing to access the data must:

⍰ You must reaffirm your commitment to the Synapse Pledge and must abide by the guiding principles for responsible research use and data handling within the Synapse Commons Platform as described in the Synapse Governance documents.
⍰ You will not attempt to establish the identity of, or attempt to contact any of the subjects included in the data.
⍰ You confirm that if you inadvertently receive identifiable information or otherwise identify a subject, you will promptly notify the ACT by emailing.
⍰ You agree to establish appropriate administrative, technical, and physical safeguards to prevent unauthorized use of or access to the Data.
⍰ You will report any data misuse or breach of data security to ACT by emailing.
⍰ You will use the data only as identified in your intended data use statement (IDU), submitted through Synapse. The IDU should be written in English and must describe the objectives of the proposed research and study design and analysis plan (500 word maximum).
⍰ Data accessors must acknowledge the following in all publications or presentations as follows: “The datasets used for the analyses described were contributed by Dr. Adithya Cattamanchi at UCSF and Dr. Simon Grandjean Lapierre at University of Montreal and were generated in collaboration with researchers at Stellenbosch University (PI Grant Theron), Walimu (PIs William Worodria and Alfred Andama); De La Salle Medical and Health Sciences Institute (PI Charles Yu), Vietnam National Tuberculosis Program (PI Nguyen Viet Nhung), Christian Medical College (PI DJ Christopher), Centre Infectiologie Charles Mérieux Madagascar (PIs Mihaja Raberahona & Rivonirina Rakotoarivelo), and Ifakara Health Institute (PIs Issa Lyimo & Omar Lweno) with funding from the U.S. National Institutes of Health (U01 AI152087), The Patrick J. McGovern Foundation and Global Health Labs.”

## ACKNOWLEDGMENT

We thank all patients and families who participated in this study. We thank healthcare providers, research personnel, laboratory technicians involved in patient recruitment and data collection.

## Funders / operations

Data from the CODA TB DREAM Challenge was generated with support from Global Health Labs, the Patrick J. McGovern Foundation, and the National Institute of Allergy and Infectious Diseases of the US National Institutes of Health under award number U01AI152087. The CODA TB DREAM Challenge and post-challenge evaluation was funded in part by the Bill & Melinda Gates Foundation.

## Direct salary support

SGL is supported by a Junior 1 Salary Award from the Fonds de Recheche Santé Québec. DJ is supported by funding by the National Institutes of Health

## AUTHOR CONTRIBUTION

Conception (SKS, LO, CB, PD, PMS, AC, SGL), data acquisition (MR, RR, IL, OL, DJC, NVN, GT, WW, CY), data analysis (SH, VY, SKS, SK, DJ), data interpretation (N/A), drafting the work (SH, SGL), reviewing the work critically for important intellectual content and final approval of the version to be published (all co-authors).

Agreement to be accountable for all aspects of the work in ensuring that questions related to the accuracy or integrity of any part of the work are appropriately investigated and resolved (SH, SKS, AC, SGL).

## COMPETING INTERESTS

The authors declare no competing interest.

## Notes

### Competing Interest Statement

The authors have declared no competing interest.

### Author Declarations

Rapid Research in Diagnostic Development TB Network (R2D2 TB Network) study: Ethical approval for this study was obtained from institutional review boards (IRB) in the US and in each study site. In the US, approval was obtained from the University of California San Francisco IRB (# 20-32670). In Vietnam, approval was obtained from the Ministry of Health Ethical Committee for National Biological Medical Research (94/CN-HDDD), the National Lung Hospital Ethical Committee for Biological Medical Research (566/2020/NCKH) and the Hanoi Department of Health, Hanoi Lung Hospital Science and Technology Initiative Committee (22/BVPHN). In India, approval was obtained from Christian Medical College IRB (13256). In South Africa, approval was obtained from Stellenbosch University Health Research Ethics Committee (17047). In Uganda, approval was obtained from Makerere University, College of Health Sciences, School of Medicine, Research Ethics Committee (2020-182). In the Philippines, approval was obtained from De La Salle Health Sciences Institute Independent Ethics Committee (2020-33-02-A). The Digital Cough Monitoring for screening, diagnosis and clinical follow-up of tuberculosis and other respiratory diseases project: Ethical approval for this study was obtained from institutional review boards (IRB) in Canada and in each study site. In Canada, approval was obtained from the Centre de Recherche du Centre Hospitalier de l Universite de Montreal IRB (# 2021-9270, 20.226). In Madagascar, approval was obtained from the Comite d Ethique a la Recherche Biomedicale (IORG0000851 - N051-MSANP/SG/AMM/CERBM).). In Tanzania, approval was obtained from the Ifakarah Health Institute IRB (31-2021) and the National Institute for Medical Research (NIMR/HQ/R.8a/Vol IX/3805).

